# Patient-keyed aggregation of public cancer genomic data imports clonal hematopoiesis into solid-tumor mutation matrices

**DOI:** 10.64898/2026.10.10.26365222

**Authors:** Lillian Zhi, Joyce Chi, Andreana Yang Leow, Minggui Pan

## Abstract

Large-scale reuse of public cancer genomic data begins by pooling studies into one patient-by-gene matrix, keyed on the patient. Records held under one patient identifier need not come from the same kind of specimen: three clonal-hematopoiesis studies in cBioPortal deposit blood normals sharing patient identifiers with tumors deposited elsewhere, so a patient-keyed merge records a hematopoietic clone as a tumor mutation. We linked 1, 282, 483 coding calls in 140 genes from 483 studies to patients, classified each by specimen type, and rebuilt the matrix with and without blood-derived calls. Blood normals supply 1.7% of calls but concentrate: 52.8% of patients flagged *DNMT3A*-mutant, 21.3% of *TET2* and 19.7% of *CHEK2* have no tumor-derived call in that gene, against 0.4% for *TP53*. Those calls carry a hematopoietic gene spectrum and, in *TP53*, hematopoietic allele fractions. The artifact reaches 65 further cohorts, almost all solid-tumor studies, exceeding one patient in ten in fifteen of them, and tracks the share of patients with a paired blood normal, not the disease (*r* = 0.994, 45 cancer types). Correction lowers the pan-cancer *DNMT3A* rate by 45% and moves survival estimates further than chance removal of the same patients would. Two filter lines applied before aggregation remove it.

## 1 Introduction

Aggregated public repositories are the substrate for a large share of computational cancer research. Using them at scale means pooling records from many separately deposited studies into a single patient-by-gene matrix, and that pooling requires a join key. The patient is the obvious choice, because the same patient genuinely does appear in several studies and consolidating those records is the point of the exercise.

Records held under one patient identifier need not, however, come from the same kind of specimen. Where they do not, and the join consults only the identifier, the resulting matrix acquires entries whose provenance differs from the provenance a downstream analysis would ordinarily assume. That provenance is not missing from the source: specimen type is recorded in the deposited files and is discarded by the join. Because the product carries no record of it, the resulting error cannot be seen in the matrix at all. It can be found only by returning to the call table.

We describe an instance of this hazard that is large, current and easily reproduced. Reuse of the many studies deposited in cBioPortal requires exactly the pooling described above, and the portal is structured so that a patient-keyed merge is the natural way to do it (Cerami et al., 2012; Gao et al., 2013; de Bruijn et al., 2023). Three Memorial Sloan Kettering clonal-hematopoiesis studies (Bolton et al., 2020; Spitzer et al., 2022; Stonestrom et al., 2024) deposit blood normals from patients whose tumors are separately deposited elsewhere, and the blood specimens carry the same patient identifier as the tumor. A merge that asks only which patient a call belongs to, and not which specimen it came from, records a hematopoietic clone as a mutation in that patient’s solid tumor.

No variant is misidentified at any point. The clonal-hematopoiesis calls were correctly called, correctly labeled, correctly published and correctly deposited. The error is created downstream, by aggregation, and it is invisible in the resulting matrix because nothing in the matrix records specimen type.

Clonal hematopoiesis is common in patients with cancer and rises steeply with age (Jaiswal et al., 2014; Genovese et al., 2014; Steensma et al., 2015; Ptashkin et al., 2018). Its capacity to masquerade as a tumor mutation is well described in the setting where it was first recognized: tumor-only sequencing without a matched normal, where a hematopoietic clone infiltrating a tumor specimen cannot be distinguished from a somatic tumor event (Ptashkin et al., 2018). That recognition is one reason paired tumor-normal sequencing became standard where it could be afforded (Zehir et al., 2017). The route described here is different, and paired sequencing does not close it. These data are paired, and it is precisely the correctly sequenced, correctly labeled blood normal that supplies the contaminating call.

Distinguishing this from a genuine biologic phenomenon also matters. Pich et al. (2025) showed that hematopoietic clones physically infiltrate solid tumors: among patients with circulating clonal hematopoiesis, the same variant was detectable in tumor tissue at a variant allele fraction of 2% or more in 42% of early-stage lung cancers and 26% of a pan-cancer cohort, with single-cell genotyping localizing the clone to tumor-associated myeloid populations rather than to admixed blood. Clonal hematopoiesis can therefore reach a tumor genotype by two quite different routes: infiltration, which is biology, and the merge described here, which is not. The two are separable by specimen, and only the second is correctable by filtering.

We quantify how far the second route reaches, show that the affected calls carry the expected clonal-hematopoiesis signature, identify the study cohorts in which the contaminated flags come to rest, demonstrate that removing them reconciles two independent measurements of the same quantity, and give the correction as a filter list and a two-line rule.

## 2 Methods

### 2.1 Data and cohort assembly

Mutation, copy-number, clinical and survival tables were extracted from 483 cBioPortal studies. Coding calls in 140 cancer-associated genes were linked to patients using the key a pooled analysis uses: MSK: <PATIENT> for Memorial Sloan Kettering patients, TCGA: <BARCODE> for The Cancer Genome Atlas, and study: <SAMPLE> otherwise. A patient-by-gene matrix was then built as a union: an entry is set if any sample belonging to that patient, in any contributing study, carried a coding call in that gene.

### 2.2 Classifying calls by specimen

The three contributing clonal-hematopoiesis studies are identified in cBioPortal as msk_ch_2020 (Bolton et al., 2020; 24, 146 patients), msk_ch_2023 (Stonestrom et al., 2024; 47, 532 samples) and msk_ch_ped_2021 (Spitzer et al., 2022). All three deposit variants called on blood specimens from patients sequenced by MSK-IMPACT with a matched normal, and it is those blood specimens, not the tumors, that carry the calls examined here.

A call was classified non-tumor if its study was msk_ch_2020, msk_ch_2023 or msk_ch_ped_2021, or if its sample identifier matched the pattern denoting a blood normal. Cell-free DNA assays were identified separately by sample identifier and were not counted as non-tumor. The corrected matrix was rebuilt from the filtered call table by the identical procedure. As a check that the correction isolates the intended calls and introduces nothing else, the same rebuild without the filter was compared cell by cell with the original matrix.

### 2.3 Locating the affected cohorts

Each contaminated gene-patient entry was attributed to the study supplying that patient’s representative tumor record, which is the cohort an investigator analyzing that study would be working in. Cohort denominators are the distinct patient identifiers in each study’s clinical sample file. Among the 65 cohorts that are not themselves clonal-hematopoiesis studies, every representative specimen was checked against that file and none is recorded as a blood normal, so the affected patients are genuine tumor cases and the contaminating call reached them only through the merge. Three of those cohorts are hematologic rather than solid-tumor studies and do not enter the solid-tumor analysis set.

### 2.4 Analysis set and statistics

The solid-tumor analysis set excludes the three clonal-hematopoiesis studies, patients whose representative record is a blood-normal, cell-line or xenograft specimen, and patients with no specimen other than blood. Frequencies are simple proportions. Cox models are Breslow-tied and adjusted for age; each gene-by-cancer-type comparison required at least 40 mutant patients, 80 deaths and 15 mutant patients after correction. Because the corrected arm necessarily has fewer mutant patients, and because losing patients moves an estimate on its own, each comparison was tested against a permutation null: from the uncorrected mutant set we removed the same number of patients the correction removed, 200 times, and recorded how often the log hazard ratio moved at least as far as correction moved it. Multiplicity within families of tests is controlled by the Benjamini-Hochberg procedure. Analyses used Python 3.11 with pandas, NumPy, SciPy and statsmodels.

## 3 Results

### 3.1 Scale of the contamination and the genes affected

Of 1, 282, 483 coding calls linked to a patient, 21, 515 (1.7%) originate in a blood normal. Every call from a clonal-hematopoiesis study sits on a blood-normal specimen identifier, and only four blood-normal calls occur outside those three studies, one of which is a pattern-matching false positive. There is no second route into the matrix.

Rebuilding the matrix from the same call table without the filter reproduces the original exactly: zero differing entries across 140 genes and 153, 264 patients. Applying the filter removes 12, 009 gene-patient entries across 9, 699 patients, 2.2% of all entries, and adds none. The corrected matrix is therefore a strict subset of the original, and the difference between them is exactly the set of blood-derived calls.

That 2.2% is not spread evenly (Fig. 1A, Table 1). More than half of all patients flagged *DNMT3A*-mutant have never had a *DNMT3A* mutation found in a tumor, and the genes at the top of the list are the canonical clonal-hematopoiesis genes. The genes an investigator is most likely to check, such as *TP53* and *KRAS*, are essentially untouched, which is why a sanity check on familiar genes passes.

**Fig. 1.**
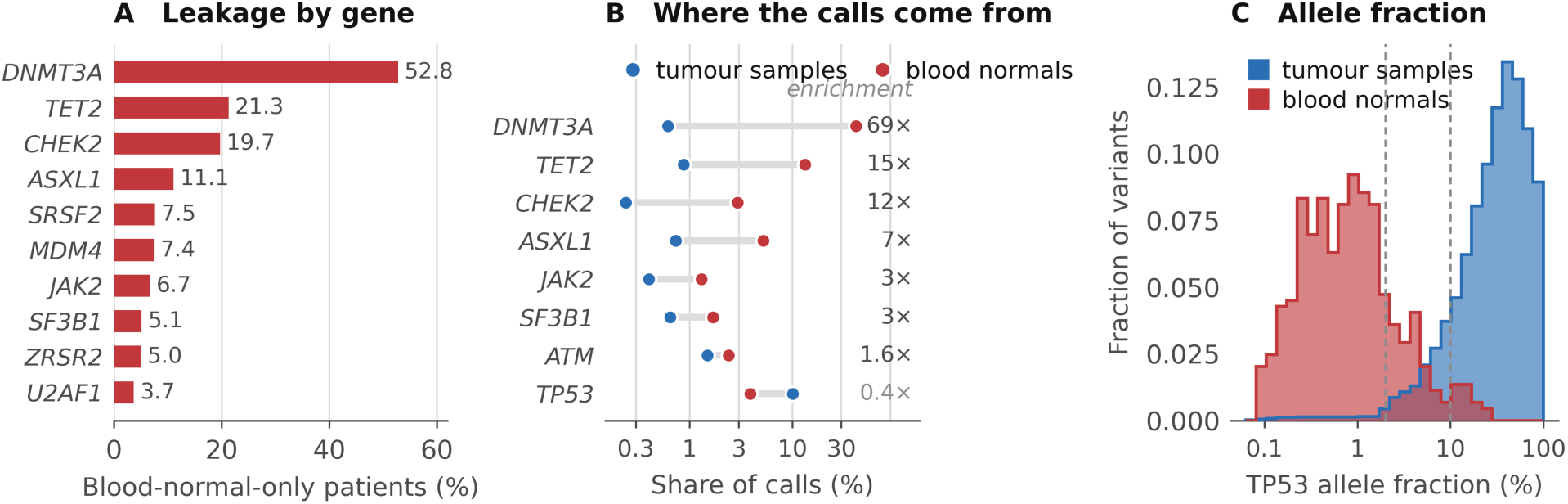
Clonal-hematopoiesis calls in the tumor gene matrix. (A) Percentage of patients flagged mutant whose only call for that gene comes from a blood-normal specimen; the ten most affected genes of 140. (B) Share of calls contributed by each gene, separately for blood-normal and tumor specimens, on a logarithmic scale. *DNMT3A* supplies 42.0% of blood-normal calls and 0.61% of tumor calls. (C) Variant allele fraction of *TP53* variants by specimen type; *TP53* is the only gene in the extract carrying read counts. Blood-normal variants have a median allele fraction of 0.4%, tumor variants 33.4%. Dashed vertical lines mark 2% and 10% allele fraction, the band in which tumor-infiltrating clonal hematopoiesis would appear.

**Table 1.** Leakage by gene.

| Gene | Patients flagged mutant | With a solid-tumor call | Only a blood-normal call, No. (%) | Only a cell-free call, No. (%) |
| --- | --- | --- | --- | --- |
| DNMT3A | 9,210 | 4,297 | 4,861 (52.8) | 52 (0.6) |
| TET2 | 7,071 | 5,560 | 1,509 (21.3) | 2 (0.0) |
| CHEK2 | 1,907 | 1,530 | 376 (19.7) | 1 (0.1) |
| ASXL1 | 5,600 | 4,977 | 622 (11.1) | 1 (0.0) |
| SRSF2 | 1,914 | 1,769 | 144 (7.5) | 1 (0.1) |
| JAK2 | 2,742 | 2,558 | 184 (6.7) | 0 (0.0) |
| SF3B1 | 4,705 | 4,344 | 242 (5.1) | 119 (2.5) |
| ATM | 7,783 | 7,271 | 283 (3.6) | 229 (2.9) |
| TP53 | 55,454 | 54,393 | 235 (0.4) | 826 (1.5) |
| KRAS | 18,582 | 18,237 | 36 (0.2) | 309 (1.7) |

### 3.2 The contaminating calls are hematopoietic clones

Three lines of evidence identify what has entered the matrix. First, the gene spectrum is not the spectrum of tumor calls (Fig. 1B). *DNMT3A* supplies 42.0% of all non-tumor calls but 0.61% of tumor calls, a 69-fold enrichment; *TET2* is enriched 15-fold, *CHEK2* 12-fold, *ASXL1* sevenfold and *SRSF2* fivefold. These are the canonical clonal-hematopoiesis genes.

Second, allele fraction. *TP53* is the only gene in this extract carrying read counts. Blood-normal *TP53* variants have a median variant allele fraction of 0.4%, with 97.1% below 10% and 85.0% below 2%, at a median depth of 698; tumor *TP53* variants have a median of 33.4% (Fig. 1C). The two distributions barely overlap. These are low-level clones detected because the clonal-hematopoiesis studies deliberately applied sensitive calling to the normal specimen, not incidental germline findings.

Third, age. *CHEK2*, *ATM* and *BRCA2* are germline predisposition genes, raising the alternative that their blood calls are germline pathogenic variants correctly reported. Germline carriage does not vary with age; acquired clones do. Among patients with a paired blood normal, the blood-only call rate for *CHEK2* rises from 1.0% under 45 years to 4.3% at 75 years and above, and for *ATM* from 0% to 3.7%. *BRCA2*, which has no clonal-hematopoiesis association, stays flat and low. Patients whose only call is a blood normal are also older than those with a tumor-derived call in the same gene: *TET2* 71.6 versus 68.0 years, *ASXL1* 72.0 versus 67.0, *CHEK2* 70.4 versus 62.0 (all *P* < .001). The age gradient establishes that these are acquired clones rather than germline variants; it is the gene spectrum and the allele fractions that identify the clones as clonal hematopoiesis, since the per-decade odds of a blood-only call rise about as steeply for *NF1* (1.58) as for *DNMT3A* (1.60).

### 3.3 Which cohorts carry the contamination

The three clonal-hematopoiesis studies are the source of the contaminating calls, but they are not where the damage is done. Of the 9, 699 affected patients, 5, 306 have their representative tumor record in one of 65 other cBioPortal study cohorts, almost all of them ordinary solid-tumor studies that deposited no blood normal of their own and whose investigators had no part in generating a clonal-hematopoiesis call. Three of the 65 are hematologic cohorts that do not enter the solid-tumor analysis set, and every representative specimen in the remaining cohorts is recorded by its own study as a tumor specimen. Two carry identifiers that match the blood-normal pattern without being blood normals: one is the pattern-matching false positive noted above, and one carries an -N01 identifier while its own study records it as a metastasis. An analysis that pools any of these cohorts with the rest of the portal inherits the flags.

The burden is concentrated (Table 2). Fifteen of the 65 cohorts exceed 10%, and eleven of the 49 that contain at least 100 patients do: the rate reaches 23.8% in the MSK lung adenocarcinoma cohort, 21.9% in an MSK esophagogastric cohort, 21.5% in the paired bladder cohort and 20.8% in the MSK gastrointestinal stromal tumor cohort. The largest affected cohort in absolute terms is MSK-CHORD, where 3, 129 of 24, 950 patients (12.5%) carry at least one contaminated flag. *DNMT3A* is the most affected gene in 52 of the 65 cohorts. The median across the 49 cohorts of at least 100 patients is 3.3%, so the problem is not universal; it is severe in a minority of cohorts, and those cohorts are not identifiable without returning to the call table.

**Table 2.** Cohorts in which more than one patient in ten carries a contaminated gene flag.

| Study | Patients | Affected, No. (%) | Contaminated entries |
| --- | --- | --- | --- |
| lung_msk_2017 | 860 | 205 (23.8) | 267 |
| egc_msk_tp53_ccr_2022 | 237 | 52 (21.9) | 57 |
| paired_bladder_2022 | 1,244 | 267 (21.5) | 321 |
| gist_msk_2022 | 499 | 104 (20.8) | 133 |
| chol_msk_2018 | 192 | 36 (18.8) | 44 |
| glioma_mskcc_2019 | 924 | 141 (15.3) | 179 |
| egc_msk_2017 | 305 | 42 (13.8) | 47 |
| msk_chord_2024 | 24,950 | 3,129 (12.5) | 3,890 |
| hcc_msk_2024 | 1,370 | 163 (11.9) | 208 |
| appendiceal_msk_2022 | 273 | 29 (10.6) | 32 |
| cervix_msk_2023 | 177 | 18 (10.2) | 25 |

Drawn from the 49 cohorts that are not themselves clonal-hematopoiesis studies and that contain at least 100 patients; eleven of those 49 exceed 10% and all eleven are shown. Four smaller cohorts also exceed 10% and are listed in the supplementary data. Patients are the distinct patient identifiers in the study's clinical sample file. An affected patient has at least one gene flag that rests only on a blood normal deposited in a different study. The complete list of all 67 affected cohorts, with the two clonal-hematopoiesis studies that supply a representative record flagged, is in the supplementary data.

### 3.4 The distortion is a property of the cohort, not the disease

Broken out by cancer type the distortion appears disease-specific and dramatic: *DNMT3A* in gastrointestinal stromal tumor 8.49% against a corrected 0.75%, in non-small-cell lung cancer 9.15% against 2.35%, in prostate cancer 5.34% against 0.75%. That reading is wrong. Across all 45 cancer types with at least 300 patients, the distortion tracks the share of that type’s patients who have a paired clonal-hematopoiesis blood normal at *r* = 0.994 (*P* = 3 × 10⁻⁴³; Fig. 2A). It is a per-study effect wearing a disease label, and any analysis that stratifies by disease will attribute a property of the sequencing cohort to biology.

**Fig. 2.**
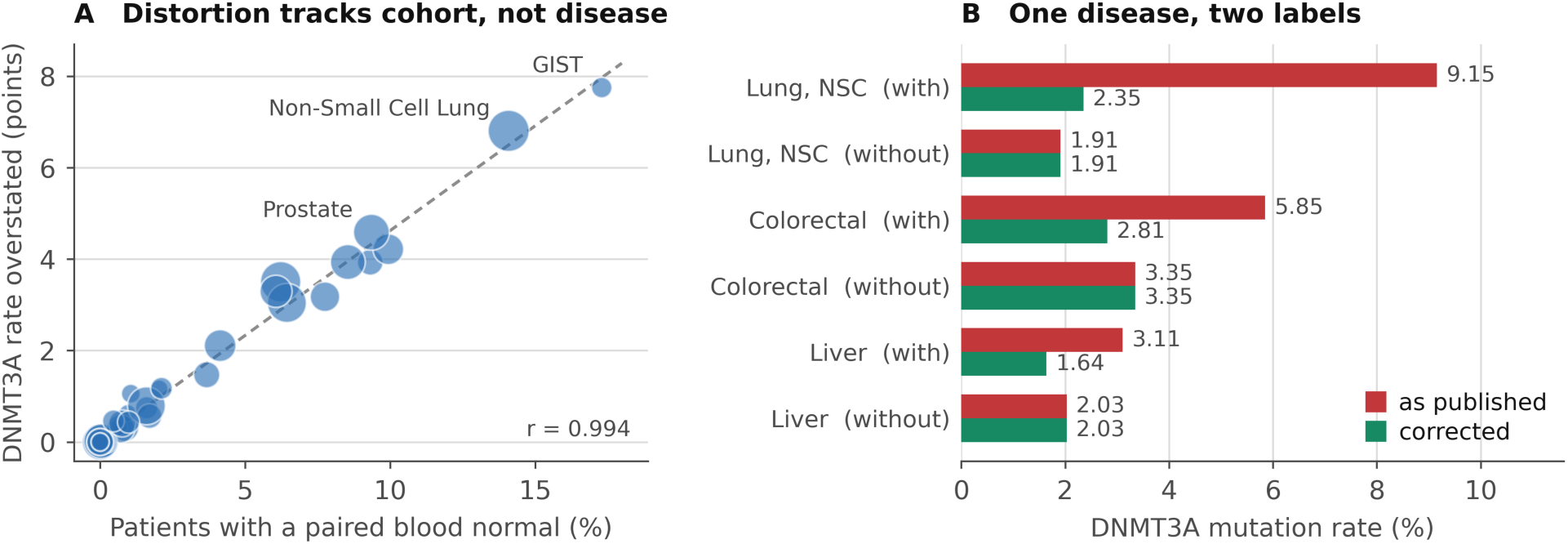
The distortion is a property of the cohort, not the disease. (A) For each of 45 cancer types with at least 300 patients, the overstatement of the *DNMT3A* mutation rate against the percentage of that type’s patients who have a paired clonal-hematopoiesis blood normal; marker area is proportional to the square root of cohort size. (B) *DNMT3A* mutation rate before and after correction for three diseases that each appear under two unharmonized cancer-type labels, one drawn from studies with paired blood normals and one without. The labels disagree by up to 4.8-fold before correction and agree within 1.3-fold after it.

The cleanest demonstration is an accident of curation. Although OncoTree provides a controlled vocabulary for tumor type (Kundra et al., 2021), the free-text cancer-type field carried alongside it in aggregated data is not harmonized, so one disease can appear under two labels drawn from different study mixes (Fig. 2B). Before correction the two labels for a single disease differ by 4.8-fold in non-small-cell lung cancer, 1.7-fold in colorectal cancer and 1.5-fold in hepatocellular carcinoma. After correction all three pairs agree within 1.3-fold and the residual differences no longer run in a consistent direction (Table 3). This is the strongest evidence that the leak is real and that removing the affected entries is the appropriate repair, because the repair reconciles two independent measurements of the same quantity that disagreed beforehand. No parameter was tuned to make them agree.

**Table 3.**
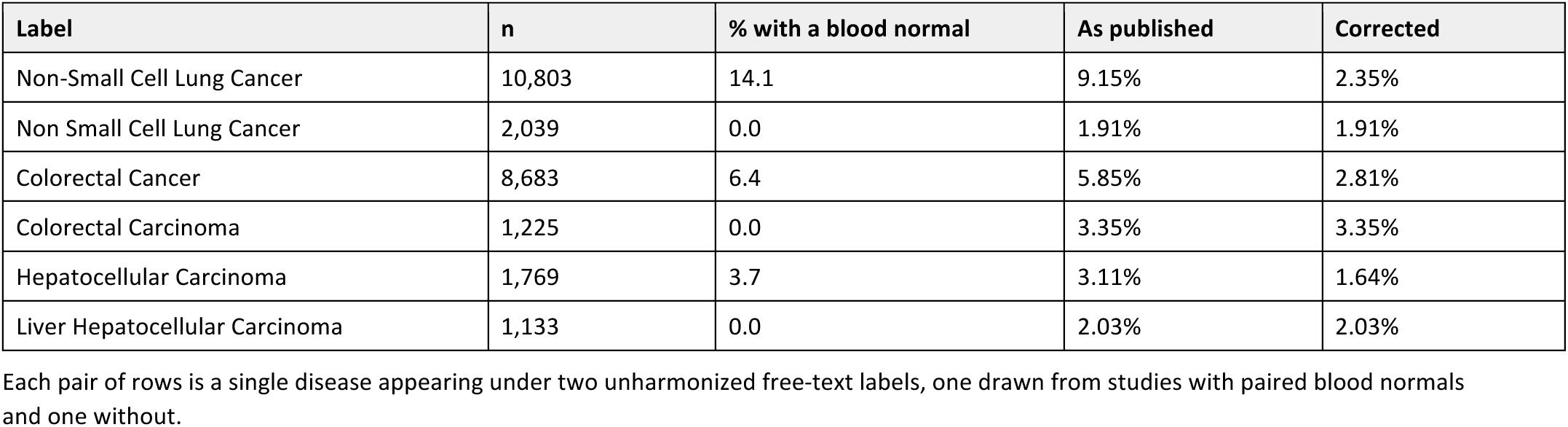
*DNMT3A* mutation rate for one disease under two labels.

| Label | n | % with a blood normal | As published | Corrected |
| --- | --- | --- | --- | --- |
| Non-Small Cell Lung Cancer | 10,803 | 14.1 | 9.15% | 2.35% |
| Non Small Cell Lung Cancer | 2,039 | 0.0 | 1.91% | 1.91% |
| Colorectal Cancer | 8,683 | 6.4 | 5.85% | 2.81% |
| Colorectal Carcinoma | 1,225 | 0.0 | 3.35% | 3.35% |
| Hepatocellular Carcinoma | 1,769 | 3.7 | 3.11% | 1.64% |
| Liver Hepatocellular Carcinoma | 1,133 | 0.0 | 2.03% | 2.03% |

Each pair of rows is a single disease appearing under two unharmonized free-text labels, one drawn from studies with paired blood normals and one without.

### 3.5 Effect on reported mutation frequencies

Across the solid-tumor analysis set the correction lowers the apparent pan-cancer *DNMT3A* mutation rate from 5.13% to 2.83%, removing 45% of its value, and lowers *TET2* by 19% and *CHEK2* by 13% (Table 4). *TP53*, *KRAS* and *PIK3CA* are unchanged to two decimal places.

**Table 4.** Pan-cancer mutation rate across 114, 141 solid-tumor patients.

| Gene | As published | Corrected | Relative overstatement |
| --- | --- | --- | --- |
| DNMT3A | 5.13% | 2.83% | 44.9% |
| TET2 | 3.92% | 3.16% | 19.3% |
| CHEK2 | 1.16% | 1.01% | 13.1% |
| ASXL1 | 3.07% | 2.74% | 10.7% |
| SRSF2 | 0.99% | 0.92% | 7.0% |
| JAK2 | 1.78% | 1.70% | 4.7% |
| SF3B1 | 2.69% | 2.57% | 4.6% |
| TP53 | 37.04% | 36.93% | 0.3% |
| KRAS | 13.25% | 13.24% | 0.1% |
| PIK3CA | 10.70% | 10.70% | 0.0% |

Relative overstatement is (published – corrected) / published, computed from the unrounded rates; recomputing it from the two-decimal values displayed here will differ slightly in the last digit. Two features of the analysis set should be borne in mind. It excludes approximately 18,000 patients whose representative record was their blood normal, precisely the population in which leakage is most concentrated, so the distortion shown here understates the effect. And both columns are attenuated by approximately 15% because 13.7% of the analysis set belongs to a study that does not report *DNMT3A* at all, which lowers both absolute percentages without affecting the ratio between them. The values are therefore a measure of distortion, not population prevalences.

### 3.6 Effect on survival estimates

Forty-eight gene-by-cancer-type comparisons met the prespecified thresholds. Thirteen moved further than random removal of the same number of patients would move them at *P* < .05, against 2.4 expected, so the correction changes survival estimates more than the loss of sample size alone accounts for. The raw count of comparisons that crossed the .05 significance threshold, seven, is not informative, because random removal produces 5.4 such changes on average.

Four comparisons both changed significance and beat the permutation null at *P* < .01. In glioma, *TET2* mutation appeared protective before correction (hazard ratio [HR], 0.58; *P* = .0004) and did not after (HR, 0.81; *P* = .22). In colorectal cancer, *DNMT3A* appeared null before correction (HR, 0.93; *P* = .52) and protective after (HR, 0.40; *P* = .0003); endometrial cancer showed the same reversal (HR, 0.78 to 0.22), as did *CHEK2* in colorectal cancer (HR, 0.60 to 0.35). The contamination therefore works in both directions: it manufactures associations and it conceals them.

These four are demonstrations that estimates move, not findings about the genes, and they should not be read as evidence about *DNMT3A*, *TET2* or *CHEK2* in those diseases. Three of the four rest on fewer than 20 deaths in the corrected mutant arm, one of them on only three. A further caution applies to all of them: simply having a paired blood normal is itself strongly associated with survival (HR 2.76 in prostate cancer, 0.68 in glioma) because it marks membership of a particular sequencing cohort. Part of what the contamination creates is a study indicator smuggled into a gene variable. Removing the affected entries is correct and necessary, but it does not by itself make either estimate free of that confounding.

There is a second and less obvious reason the contaminating variable is prognostically loaded. Clonal hematopoiesis is not acquired at random with respect to treatment: DNA-damage-response clones carrying *TP53*, *PPM1D* and *CHEK2* mutations are selectively expanded by radiation, platinum agents and topoisomerase II inhibitors, and *ASXL1* clones are enriched in smokers (Bolton et al., 2020). The leaked calls are therefore concentrated in patients who have had more prior cytotoxic therapy, which is itself a marker of more advanced or more heavily treated disease. A contaminating exposure that tracks treatment history will not behave like random misclassification, which attenuates toward the null; it can bias an estimate in either direction, and it does so most strongly for exactly the genes most affected here. *CHEK2*, at 19.7% blood-normal-only, is both a therapy-selected clonal-hematopoiesis gene and a gene whose germline variants are studied for their effect on outcome, which makes it the single most hazardous entry in Table 1 to leave uncorrected.

## 4 Implementation

The correction is two filters applied to the call table before patient-level aggregation, not to the matrix afterward: exclude calls from the three clonal-hematopoiesis studies, and exclude calls whose specimen identifier denotes a blood normal.

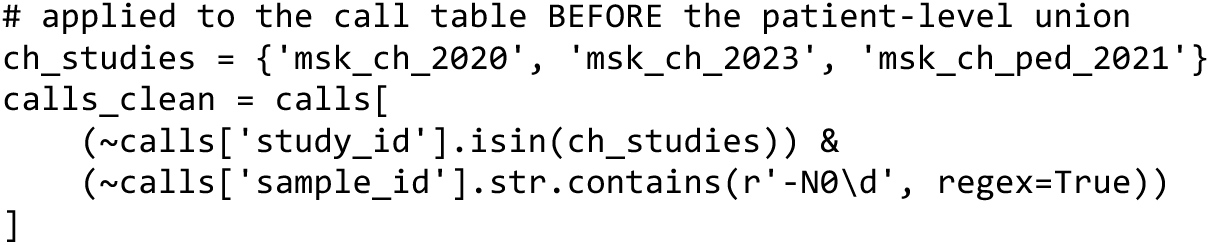

The sample-identifier pattern matters. MSK blood normals are numbered specimens of the form P-0000000-N01, so the pattern must admit the digits; a pattern anchored on a bare -N suffix matches 2 of the 21, 515 affected calls in these data and silently leaves the artifact in place. Any implementation should be checked by counting what it removes before it is trusted.

For analyses already completed, the supplementary data list all 12, 009 affected gene-patient entries, so they can be removed from an existing matrix without rebuilding it. A standalone validation script reproduces the per-gene leakage measurement reported here directly from the public data; its test for a non-tumor specimen is deliberately broader than the rule above, keying on the sample-type and sample-class fields as well as the identifier pattern, so that it remains valid on snapshots in which the identifier convention differs. Both are described under Software and Data availability.

Two nuances warrant attention in any implementation. First, cell-free DNA assays are a separate and smaller route into pooled matrices, affecting 2.9% of *ATM*-flagged, 2.5% of *SF3B1*-flagged and 1.5% of *TP53*-flagged patients (Table 1); their spectrum is not clonal-hematopoiesis-enriched, but multi-analyte workflows should partition cell-free specimens into a separate matrix rather than pooling them with solid-tumor tissue calls. Second, a patient with a genuine tumor mutation retains the entry even if they also carry a blood clone in the same gene, which is the conservative choice.

## 5 Discussion

A patient-keyed merge across public cancer genomic studies records clonal hematopoiesis as tumor genotype. The affected genes are precisely those a hematology-adjacent or aging-related analysis would want to study; the distortion is large enough to move a pan-cancer *DNMT3A* frequency by 45% of its value and to reverse the direction of survival associations; and it reaches 65 other study cohorts, almost all of them solid-tumor studies whose own investigators deposited no blood normal.

This is neither of the two failure modes already described. It is not the tumor-only sequencing problem (Ptashkin et al., 2018), and it is not tumor-infiltrating clonal hematopoiesis (Pich et al., 2025). Pich et al. begin from a clone identified in blood and ask whether it is present in the tumor specimen; they require the two specimens to be held apart, and had they been merged on patient identity there would have been no contrast left to measure. The artifact described here is what happens when that separation is not maintained. These data are paired. The clonal-hematopoiesis calls were identified as such by the investigators who generated them, published as such, and deposited as such. Nothing is misread. The artifact is created by the join, and it will be reproduced by any pipeline that pools studies on patient identity, a common and otherwise sensible design.

Three features make it easy to miss. The overall magnitude is small: 1.7% of calls, 2.2% of entries. The genes an investigator is most likely to be checking are essentially unaffected, so a sanity check on familiar genes passes. And the resulting matrix carries no record of specimen type, so the contamination cannot be seen in the product; it can only be seen by returning to the call table.

The strongest evidence that the repair is the right one is that it reconciles measurements that disagreed before it was applied. Three diseases each appear in these data under two unharmonized labels, one drawn from cohorts with paired blood normals and one without. The two labels disagree by up to 4.8-fold on the *DNMT3A* rate before correction and agree within 1.3-fold after it. Nothing in the filter was fitted to produce that outcome; the filter operates on specimen type alone and is blind to cancer-type labels.

The most important limitation is what the correction cannot reach. It removes calls that arise in blood specimens; it cannot remove hematopoietic clones present within tumor specimens, and Pich et al. (2025) establish that those exist at appreciable frequency. The corrected rates in Table 4 are therefore upper bounds on the somatic tumor mutation rate, not estimates of it, and the residual is unlikely to be negligible for exactly the genes most affected here. In our own data 5.6% of *TP53* variants called in tumor specimens fall in the 2-10% allele-fraction band where tumor-infiltrating clonal hematopoiesis would appear, and 13.5% fall below 10%; but that fraction is no higher in patients who have a paired blood normal than in those who do not (13.8% v 13.4%; *P* = .18), so low tumor purity and subclonality account for it just as well. Separating the two requires single-cell or cell-type-resolved genotyping of the kind Pich et al. performed, which aggregated repository data cannot supply. This report corrects a merge error; it does not deliver a clean somatic tumor genotype.

Other limitations. Allele fractions were available for *TP53* only, and only from one of the three contributing studies, so for other genes the clonal-hematopoiesis interpretation rests on the gene spectrum and the age gradient rather than on direct observation of a low-level clone. The germline comparison uses small numbers at the young end. The survival analyses illustrate instability rather than establishing anything about the genes involved, and having a paired blood normal is itself a cohort marker associated with survival that the correction does not remove. *PPM1D*, the archetypal therapy-related clonal-hematopoiesis gene and among the most strongly therapy-selected of them (Bolton et al., 2020), together with *CBL* and *GNB1*, is absent from the 140-gene panel used here and is never reported as absent; because *PPM1D* expands under precisely the exposures that bring patients into these cohorts, it is the gene we would expect to head the list on a complete gene set. A re-analysis over a complete gene set would very likely identify further affected genes and would refine the per-gene figures, and the accompanying validation script is written so that it can be run directly against a full cBioPortal snapshot. Finally, the analysis rests on one snapshot of one aggregator; the specific studies and identifiers will change as cBioPortal grows. The mechanism, however, does not depend on the gene set or the snapshot: it is a property of the join.

The specific artifact described here is narrow and easily fixed. The general point is not. When records from specimens of different kinds are merged on a shared identifier, the identifier must not be the only thing the merge consults. A pooled matrix should carry the provenance of every entry; specimen type is present in the source files and discarded by the join, and nothing in the matrix itself records it. The assembly step in reuse of public genomic data is rarely reported in enough detail to be checked, and it is the step at which this class of error is introduced. That is worth saying plainly in a field now asking a genuinely interesting question about what hematopoietic clones are doing inside tumors (Pich et al., 2025), because the answer will be read out of these same repositories, and an artifact that mimics the phenomenon is a poor foundation for studying it.

## Data Availability

All data produced in the present work are contained in the manuscript

## Author contributions

Conception and design: MP. Collection and assembly of data: LZ, JC, AL, and MP. Data analysis and interpretation: LZ, JC, AL, and MP. Manuscript writing: all authors. Final approval of manuscript: all authors. Accountable for all aspects of the work: all authors.

## Declaration of generative artificial intelligence use

The authors used a generative artificial intelligence assistant (Claude, Anthropic) in preparing this work. Because that use went beyond language editing, it is described in full. Working under author direction, the assistant was used to write the analysis code — cohort assembly, classification of calls by specimen type, the Cox models and the permutation procedure — to render the figures from that code, to assemble the tables from the analysis output, and to draft and revise the text. Both figures are data plots produced by matplotlib from the analysis output; no image in this report was produced by a generative model, and no image was altered after rendering.

The study question, the analytic design, the interpretation of the findings and every claim advanced here are the authors’ own. All authors reviewed the code, the analysis output and the text, and checked each quantitative statement against its source table; errors identified in that review were corrected before submission. The authors take full responsibility for the accuracy and integrity of the work, including the parts prepared with the assistance described above. The assistant does not meet the criteria for authorship, is not an author, and cannot be accountable for the work.

Nothing in this report depends on that assistance being trusted: the validation script regenerates the central measurements from the public data independently of the code that produced them in the first instance.

## Conflict of interest

The authors declare no conflict of interest.

## Funding

No specific funding was received for this work.

## Software and Data availability

All data analyzed here are publicly available from cBioPortal (https://www.cbioportal.org). No new sequencing was performed, and this report contributes no individual-level data beyond what is already in the public domain. The analysis drew on 483 studies; the three clonal-hematopoiesis studies central to it are deposited as msk_ch_2020, msk_ch_2023 and msk_ch_ped_2021, whose primary publications are Bolton et al. (2020), Stonestrom et al. (2024) and Spitzer et al. (2022). Any further access to the underlying MSK-IMPACT data is governed by the terms of the contributing studies rather than by the present authors.

Analysis code, the complete filter list and the validation script are freely available at [GITHUB URL TO BE INSERTED] under the MIT License and archived at [ZENODO DOI TO BE INSERTED]. The repository contains validate_ch_leakage.py, a standalone script that depends only on pandas and NumPy, plus requests for the application programming interface path, together with test data, documentation and the tab-delimited file CH_flags_to_drop.tsv listing all 12, 009 affected gene-patient entries. Every field in that file is reproduced from the public deposits and contains nothing that is not already retrievable from cBioPortal. Analyses were performed in Python 3.11 with pandas, NumPy, SciPy and statsmodels. Together with the public data, the repository is sufficient to reproduce the central measurements, and no additional code, restricted resource or author-mediated access is required.

Supplementary data comprise eight tables: the 140-gene list (S0); leakage by gene for the 18 most affected genes (S1); cohort composition by cancer type (S2); the two-label comparison (S3); the complete list of all 67 affected study cohorts with denominators, with the clonal-hematopoiesis studies flagged (S3b); the germline age-gradient test (S4); all 48 gene-by-cancer-type survival comparisons with their permutation-null results (S5); and the complete list of 12, 009 affected gene-patient entries (S6).

## Notes

### Competing Interest Statement

The authors have declared no competing interest.

### Summary of Updates

We have revised the manuscript and submitted it to a different journal for review.

