## Supplementary Tables 1-8 for "Patient-keyed aggregation of public cancer genomic data imports clonal hematopoiesis into solid-tumor mutation matrices" (from another version of this preprint: version 1, 2026-10-12)

### Supplementary Data

#### More Than Half of DNMT3A-Mutant Patients in Pooled Public Cancer Genomic Data Carry No Tumor-Derived Mutation: Clonal Hematopoiesis Introduced by Patient-Keyed Aggregation

Lillian Zhi, Joyce Chi, Andreana Yang Leow, Minggui Pan

#### Contents


| | | |
|---|---|---|
| **S1** | The 140-gene panel analyzed | 140 genes |
| **S2** | Leakage by gene, the 18 most affected genes | 18 rows |
| **S3** | Germline test: blood-only call rate by age band | 4 genes |
| **S4** | Every affected study cohort, with denominators | 67 rows |
| **S5** | Cohort composition by cancer type | 12 rows |
| **S6** | One disease under two unharmonized labels | 6 rows |
| **S7** | All 48 gene-by-cancer-type survival comparisons | 48 rows |
| **S8** | Every affected gene-patient entry | 12,009 rows |


S8 is additionally supplied as a tab-delimited file, `CH_flags_to_drop.tsv`, so that investigators who have already built a pooled matrix can set those entries to zero without rebuilding it. S4 is supplied as `CH_landing_studies.tsv`. The validation script `validate_ch_leakage.py` accompanies this file and reproduces the per-gene leakage measurement against any cBioPortal snapshot.

#### Supplementary Table S1. The 140-gene panel

Coding calls in these genes were linked to patients and classified by specimen type.

| | | | | | | |
|---|---|---|---|---|---|---|
| ACVR2A | AKT1 | ALK | APC | AR | ARID1A | ARID1B |
| ARID2 | ASXL1 | ATM | ATR | ATRX | AXIN1 | AXIN2 |
| B2M | BAP1 | BCOR | BRAF | BRCA1 | BRCA2 | CALR |
| CCND1 | CCNE1 | CDH1 | CDK4 | CDK6 | CDKN1A | CDKN1B |
| CDKN2A | CEBPA | CHD4 | CHEK2 | CIC | CREBBP | CTNNB1 |
| CUL3 | DAXX | DNMT3A | EGFR | EP300 | ERBB2 | ERBB3 |
| ERBB4 | ERCC2 | ESR1 | EZH2 | FANCA | FBXW7 | FGFR1 |
| FGFR2 | FGFR3 | FGFR4 | FLT3 | FOXA1 | FUBP1 | GATA3 |
| GNA11 | GNAQ | GNAS | HRAS | IDH1 | IDH2 | JAK1 |
| JAK2 | KDM6A | KEAP1 | KIT | KMT2A | KMT2B | KMT2C |
| KMT2D | KRAS | MAP2K1 | MAP3K1 | MDM2 | MDM4 | MEN1 |
| MET | MGA | MLH1 | MSH2 | MSH6 | MTOR | MYB |
| MYC | MYCN | NCOR1 | NF1 | NF2 | NFE2L2 | NOTCH1 |
| NOTCH2 | NOTCH3 | NPM1 | NRAS | NSD1 | PALB2 | PBRM1 |
| PDGFRA | PIK3CA | PIK3R1 | PMS2 | POLD1 | POLE | PPP2R1A |
| PTEN | PTPRD | PTPRT | RAD51C | RAD51D | RASA1 | RB1 |
| RET | RHOA | RNF43 | ROS1 | RUNX1 | SETD2 | SF3B1 |
| SMAD2 | SMAD4 | SMARCA4 | SMARCB1 | SOX2 | SPOP | SRSF2 |
| STAT3 | STK11 | TCF7L2 | TERT | TET2 | TGFBR2 | TP53 |
| TSC1 | TSC2 | U2AF1 | VHL | WT1 | ZFHX3 | ZRSR2 |

#### Supplementary Table S2. Leakage by gene

The 18 most affected genes of the 140 analyzed. "Only a blood-normal call" counts patients flagged mutant in that gene whose sole evidence is a blood-normal specimen.

| gene | patients flagged mutant | with a solid-tumour call | only a blood-normal call | % blood-normal only | only a cell-free call | % cell-free only |
|---|---|---|---|---|---|---|
| DNMT3A | 9,210 | 4,297 | 4,861 | 52.8 | 52 | 0.56 |
| TET2 | 7,071 | 5,560 | 1,509 | 21.3 | 2 | 0.03 |
| CHEK2 | 1,907 | 1,530 | 376 | 19.7 | 1 | 0.05 |
| ASXL1 | 5,600 | 4,977 | 622 | 11.1 | 1 | 0.02 |
| SRSF2 | 1,914 | 1,769 | 144 | 7.5 | 1 | 0.05 |
| MDM4 | 704 | 652 | 52 | 7.4 | 0 | 0 |
| JAK2 | 2,742 | 2,558 | 184 | 6.7 | 0 | 0 |
| SF3B1 | 4,705 | 4,344 | 242 | 5.1 | 119 | 2.53 |
| ZRSR2 | 1,065 | 1,012 | 53 | 5 | 0 | 0 |
| U2AF1 | 1,743 | 1,663 | 65 | 3.7 | 15 | 0.86 |
| GNAS | 2,524 | 2,395 | 92 | 3.6 | 37 | 1.47 |
| ATM | 7,783 | 7,271 | 283 | 3.6 | 229 | 2.94 |
| CUL3 | 1,549 | 1,508 | 41 | 2.6 | 0 | 0 |
| CALR | 783 | 762 | 20 | 2.6 | 1 | 0.13 |
| STAT3 | 1,519 | 1,474 | 39 | 2.6 | 6 | 0.39 |
| EZH2 | 2,233 | 2,175 | 55 | 2.5 | 3 | 0.13 |
| IDH2 | 1,791 | 1,740 | 42 | 2.3 | 9 | 0.5 |
| MYCN | 1,283 | 1,249 | 23 | 1.8 | 11 | 0.86 |

#### Supplementary Table S3. Germline test

Blood-only call rate (%) by age band among patients with a paired blood normal. Germline carriage does not vary with age; acquired clones do. BRCA2 has no clonal-hematopoiesis association and is the negative control.

| gene | under 45 | 45-55 | 55-65 | 65-75 | 75+ |
|---|---|---|---|---|---|
| CHEK2 | 1.03% (1) | 1.16% (3) | 2.86% (20) | 3.40% (40) | 4.32% (35) |
| ATM | 0.00% (0) | 1.16% (3) | 2.00% (14) | 2.81% (33) | 3.70% (30) |
| BRCA2 | 0.00% (0) | 0.00% (0) | 0.72% (5) | 0.60% (7) | 0.86% (7) |
| DNMT3A | 45.36% (44) | 53.88% (139) | 54.36% (380) | 50.13% (589) | 40.86% (331) |

#### Supplementary Table S4. Every affected study cohort

Every cBioPortal study cohort containing at least one patient whose gene flag rests only on a blood normal deposited in a different study.

| cBioPortal study | patients in study | patients with ≥1 contaminated flag | % of study affected | contaminated gene-patient entries | most affected gene | is itself a CH study |
|---|---|---|---|---|---|---|
| msk_chord_2024 | 24,950 | 3,129 | 12.5 | 3,890 | DNMT3A | |
| pancan_mimsi_msk_2024 | 5,020 | 314 | 6.3 | 388 | DNMT3A | |
| paired_bladder_2022 | 1,244 | 267 | 21.5 | 321 | DNMT3A | |
| lung_msk_2017 | 860 | 205 | 23.8 | 267 | DNMT3A | |
| hcc_msk_2024 | 1,370 | 163 | 11.9 | 208 | DNMT3A | |
| sarcoma_mskcc_2022 | 2,138 | 145 | 6.8 | 175 | DNMT3A | |
| glioma_mskcc_2019 | 924 | 141 | 15.3 | 179 | DNMT3A | |
| gist_msk_2022 | 499 | 104 | 20.8 | 133 | DNMT3A | |
| tmb_mskcc_2018 | 1,661 | 64 | 3.9 | 79 | DNMT3A | |
| prad_msk_stopsack_2021 | 2,069 | 60 | 2.9 | 76 | DNMT3A | |
| pdac_msk_2024 | 2,336 | 55 | 2.4 | 68 | DNMT3A | |
| egc_msk_tp53_ccr_2022 | 237 | 52 | 21.9 | 57 | DNMT3A | |
| egc_msk_2023 | 902 | 48 | 5.3 | 56 | DNMT3A | |
| nsclc_ctdx_msk_2022 | 1,127 | 43 | 3.8 | 55 | DNMT3A | |
| egc_msk_2017 | 305 | 42 | 13.8 | 47 | DNMT3A | |
| egc_mskcc_2020 | 487 | 40 | 8.2 | 49 | DNMT3A | |
| chol_msk_2018 | 192 | 36 | 18.8 | 44 | DNMT3A | |
| appendiceal_msk_2022 | 273 | 29 | 10.6 | 32 | DNMT3A | |
| breast_msk_2018 | 1,756 | 27 | 1.5 | 38 | DNMT3A | |
| csf_msk_2024 | 711 | 25 | 3.5 | 28 | DNMT3A | |
| mel_mskimpact_2020 | 696 | 23 | 3.3 | 31 | DNMT3A | |
| prad_pik3r1_msk_2021 | 1,417 | 23 | 1.6 | 27 | DNMT3A | |
| gbc_mskcc_2022 | 233 | 22 | 9.4 | 22 | DNMT3A | |
| msk_access_2021 | 617 | 20 | 3.2 | 24 | DNMT3A | |
| cervix_msk_2023 | 177 | 18 | 10.2 | 25 | DNMT3A | |
| hcc_jcopo_msk_2023 | 51 | 16 | 31.4 | 21 | DNMT3A | |
| bladder_msk_2023 | 400 | 15 | 3.8 | 23 | DNMT3A | |
| ptad_msk_2024 | 92 | 12 | 13 | 17 | TET2 | |
| crc_eo_2020 | 1,516 | 11 | 0.7 | 14 | DNMT3A | |
| luad_mskcc_2023_met_organotropism | 2,298 | 11 | 0.5 | 13 | DNMT3A | |
| bowel_colitis_msk_2022 | 175 | 11 | 6.3 | 16 | DNMT3A | |
| prostate_msk_2024 | 2,257 | 11 | 0.5 | 13 | DNMT3A | |
| gist_msk_2023 | 469 | 10 | 2.1 | 13 | TET2 | |
| gct_msk_2016 | 180 | 9 | 5 | 10 | DNMT3A | |
| thyroid_mskcc_2016 | 117 | 8 | 6.8 | 14 | DNMT3A | |
| es_dsrct_msk_2023 | 277 | 8 | 2.9 | 10 | DNMT3A | |
| sarcoma_msk_2023 | 82 | 8 | 9.8 | 10 | DNMT3A | |
| ucec_msk_2024 | 33 | 7 | 21.2 | 7 | DNMT3A | |
| crc_dd_2022 | 47 | 7 | 14.9 | 9 | DNMT3A | |
| hnc_mskcc_2016 | 151 | 6 | 4 | 7 | DNMT3A | |
| luad_mskcc_2020 | 604 | 6 | 1 | 9 | DNMT3A | |
| acc_2019 | 1,045 | 5 | 0.5 | 6 | DNMT3A | |
| prad_mskcc_2017 | 451 | 5 | 1.1 | 8 | DNMT3A | |
| breast_alpelisib_2020 | 51 | 4 | 7.8 | 4 | DNMT3A | |
| pancreas_msk_2024 | 395 | 4 | 1 | 6 | DNMT3A | |
| luad_mskimpact_2021 | 93 | 4 | 4.3 | 6 | DNMT3A | |
| ihch_mskcc_2020 | 219 | 4 | 1.8 | 4 | DNMT3A | |
| gbc_msk_2018 | 101 | 3 | 3 | 3 | RASA1 | |

| cBioPortal study | patients in study | patients with ≥1 contaminated flag | % of study affected | contaminated gene-patient entries | most affected gene | is itself a CH study |
|---|---|---|---|---|---|---|
| hgsoc_msk_2021 | 45 | 3 | 6.7 | 4 | ATM | |
| msk_spectrum_tme_2022 | 42 | 3 | 7.1 | 3 | DNMT3A | |
| difg_msk_2023 | 73 | 2 | 2.7 | 3 | DNMT3A | |
| mtnn_msk_2022 | 132 | 2 | 1.5 | 2 | ASXL1 | |
| panet_msk_2018 | 80 | 2 | 2.5 | 2 | EGFR | |
| breast_ink4_msk_2021 | 1,116 | 2 | 0.2 | 2 | PDGFRA | |
| ilc_msk_2023 | 24 | 2 | 8.3 | 2 | DNMT3A | |
| usarc_msk_2020 | 107 | 1 | 0.9 | 1 | DNMT3A | |
| blca_plasmacytoid_mskcc_2016 | 31 | 1 | 3.2 | 3 | ATM | |
| prad_cdk12_mskcc_2020 | 1,465 | 1 | 0.1 | 1 | ASXL1 | |
| mbn_msk_2024 | 363 | 1 | 0.3 | 1 | DNMT3A | |
| mds_mskcc_2020 | 4,231 | 1 | 0 | 1 | TET2 | |
| lung_msk_mind_2020 | 247 | 1 | 0.4 | 1 | MGA | |
| brca_mapk_hp_msk_2021 | 145 | 1 | 0.7 | 1 | DNMT3A | |
| panet_msk_erc_2023 | 25 | 1 | 4 | 1 | DNMT3A | |
| mixed_impact_subset_2022 | 69 | 1 | 1.4 | 1 | ASXL1 | |
| egc_trap_msk_2020 | 37 | 1 | 2.7 | 1 | TET2 | |
| msk_ch_2020 | 24,146 | 2,639 | 10.9 | 3,238 | DNMT3A | yes |
| msk_ch_2023 | 42,714 | 1,754 | 4.1 | 2,179 | DNMT3A | yes |

S4 lists every cBioPortal study cohort containing at least one patient whose gene flag rests only on a blood normal deposited in a different study. A study is attributed the patients whose representative tumour record it supplies; that is the cohort an investigator analysing that study would be working in. Denominators are distinct PATIENT_ID values in each study's data_clinical_sample.txt from the August 2026 cBioPortal bulk download. Among the 65 cohorts that are not themselves clonal-haematopoiesis studies, every representative specimen was checked against that file and none is recorded as a blood normal, so these are genuine tumour cases. Two carry identifiers matching the blood-normal pattern without being blood normals: XRN-N03JPF (acc_2019), the pattern-matching false positive, and P-0025907-N01-IM6 (mel_mskimpact_2020), which its own study records as a metastasis. Three of the 65 are haematologic rather than solid-tumour cohorts (mds_mskcc_2020, mbn_msk_2024, mtnn_msk_2022) and do not enter the solid-tumour analysis set. Two clonal-haematopoiesis studies appear because they supply a representative record for some patients; msk_ch_ped_2021 supplies none and is absent. All three are excluded from the solid-tumour analysis set. Totals: 65 non-CH cohorts, 5,306 affected patients, 6,592 contaminated entries; with the two CH studies listed, 67 cohorts and 9,699 patients.

#### Supplementary Table S5. Cohort composition by cancer type

Cancer types with at least 300 patients, ordered by the share of patients carrying a paired clonal-hematopoiesis blood normal.

| cancer type | n | % with a CH blood normal | DNMT3A as published | DNMT3A corrected | points overstated |
|---|---|---|---|---|---|
| Gastrointestinal Stromal Tumor | 671 | 17.3 | 8.49% | 0.75% | 7.75 |
| Non-Small Cell Lung Cancer | 10,803 | 14.1 | 9.15% | 2.35% | 6.8 |
| Prostate Cancer | 6,235 | 9.4 | 5.34% | 0.75% | 4.59 |
| Bladder Cancer | 3,322 | 9.9 | 7.41% | 3.19% | 4.21 |
| Pancreatic Cancer | 5,540 | 8.5 | 4.66% | 0.72% | 3.94 |
| Biliary Tract Cancer | 1,654 | 9.3 | 5.08% | 1.15% | 3.93 |
| Breast Cancer | 9,932 | 6.2 | 4.53% | 1.03% | 3.5 |
| Endometrial Cancer | 3,976 | 6.1 | 6.11% | 2.82% | 3.29 |
| Esophagogastric Cancer | 2,865 | 7.7 | 4.82% | 1.64% | 3.18 |
| Colorectal Cancer | 8,683 | 6.4 | 5.85% | 2.81% | 3.04 |
| Glioma | 3,844 | 4.1 | 4.01% | 1.90% | 2.11 |
| Hepatocellular Carcinoma | 1,769 | 3.7 | 3.11% | 1.64% | 1.47 |

#### Supplementary Table S6. One disease under two unharmonized labels

Each pair of rows is a single disease appearing under two free-text cancer-type labels, one drawn from studies with paired blood normals and one without.

| cancer_type label | n | % with a CH blood normal | DNMT3A as published | DNMT3A corrected |
|---|---|---|---|---|
| Non-Small Cell Lung Cancer | 10,803 | 14.1% | 9.15% | 2.35% |
| Non Small Cell Lung Cancer | 2,039 | 0.0% | 1.91% | 1.91% |
| Colorectal Cancer | 8,683 | 6.4% | 5.85% | 2.81% |
| Colorectal Carcinoma | 1,225 | 0.0% | 3.35% | 3.35% |
| Hepatocellular Carcinoma | 1,769 | 3.7% | 3.11% | 1.64% |
| Liver Hepatocellular Carcinoma | 1,133 | 0.0% | 2.03% | 2.03% |

#### Supplementary Table S7. All 48 gene-by-cancer-type survival comparisons

Columns 3 to 6 are the uncorrected analysis, columns 7 to 10 the corrected one. "P vs random removal" is the permutation test described in the Methods: the probability that removing the same number of mutant patients at random moves the log hazard ratio at least as far as correction moved it.

| gene | cancer type | n mut | deaths | HR | P | n mut (corr) | deaths (corr) | HR (corr) | P (corr) | HR shift | P vs random removal | chance of a flip |
|---|---|---|---|---|---|---|---|---|---|---|---|---|
| DNMT3A | Endometrial Cancer | 96 | 25 | 0.78 | 0.24 | 47 | 3 | 0.22 | 0.0091 | -0.56 | 0.005 | 0.06 |
| DNMT3A | Colorectal Cancer | 195 | 88 | 0.93 | 0.52 | 76 | 16 | 0.4 | 0.0003 | -0.53 | 0.005 | 0.03 |
| TET2 | Endometrial Cancer | 80 | 8 | 0.29 | 0.0006 | 76 | 5 | 0.19 | 0.0003 | -0.1 | 0.005 | 0 |
| TET2 | Colorectal Cancer | 150 | 51 | 0.71 | 0.02 | 105 | 26 | 0.51 | 0.0007 | -0.2 | 0.005 | 0.52 |
| TET2 | Glioma | 73 | 46 | 0.58 | 0.0004 | 53 | 36 | 0.81 | 0.22 | 0.23 | 0.005 | 0.02 |
| CHEK2 | Colorectal Cancer | 48 | 14 | 0.6 | 0.06 | 41 | 7 | 0.35 | 0.0053 | -0.26 | 0.005 | 0.26 |
| ASXL1 | Non-Small Cell Lung Cancer | 176 | 104 | 1.12 | 0.27 | 105 | 52 | 0.92 | 0.55 | -0.2 | 0.01 | 0.04 |
| TET2 | Melanoma | 51 | 27 | 0.89 | 0.54 | 48 | 27 | 0.99 | 0.97 | 0.11 | 0.01 | 0 |
| CHEK2 | Non-Small Cell Lung Cancer | 74 | 38 | 1.18 | 0.32 | 54 | 22 | 0.92 | 0.68 | -0.26 | 0.02 | 0.01 |
| TET2 | Breast Cancer | 77 | 21 | 0.96 | 0.87 | 32 | 5 | 0.52 | 0.15 | -0.44 | 0.03 | 0.01 |
| SF3B1 | Colorectal Cancer | 91 | 22 | 0.46 | 0.0003 | 85 | 18 | 0.4 | 0.0001 | -0.06 | 0.03 | 0 |
| DNMT3A | Esophagogastric Cancer | 65 | 40 | 0.82 | 0.22 | 18 | 6 | 0.48 | 0.07 | -0.34 | 0.04 | 0.05 |
| ASXL1 | Esophagogastric Cancer | 57 | 32 | 1.09 | 0.62 | 47 | 22 | 0.89 | 0.58 | -0.2 | 0.04 | 0 |
| CHEK2 | Bladder Cancer | 46 | 19 | 0.7 | 0.12 | 41 | 15 | 0.6 | 0.05 | -0.1 | 0.05 | 0.02 |
| ATM | Bladder Cancer | 125 | 46 | 0.77 | 0.09 | 124 | 45 | 0.75 | 0.07 | -0.02 | 0.05 | 0 |
| DNMT3A | Bladder Cancer | 99 | 41 | 0.69 | 0.02 | 38 | 10 | 0.47 | 0.02 | -0.21 | 0.07 | 0.8 |
| ASXL1 | Colorectal Cancer | 153 | 53 | 0.71 | 0.02 | 141 | 45 | 0.66 | 0.0068 | -0.05 | 0.07 | 0.12 |
| CHEK2 | Endometrial Cancer | 54 | 9 | 0.63 | 0.18 | 50 | 7 | 0.52 | 0.09 | -0.11 | 0.09 | 0.01 |
| SF3B1 | Endometrial Cancer | 71 | 3 | 0.12 | 0.0003 | 69 | 3 | 0.13 | 0.0004 | 0.01 | 0.1 | 0 |
| ATM | Esophagogastric Cancer | 90 | 32 | 0.58 | 0.0028 | 86 | 28 | 0.54 | 0.0016 | -0.04 | 0.11 | 0 |
| ATM | Non-Small Cell Lung Cancer | 312 | 183 | 1.16 | 0.05 | 293 | 168 | 1.13 | 0.13 | -0.03 | 0.13 | 0.34 |
| ATM | Endometrial Cancer | 155 | 14 | 0.28 | 0 | 154 | 14 | 0.28 | 0 | 0 | 0.15 | 0 |
| ASXL1 | Pancreatic Cancer | 41 | 29 | 0.74 | 0.1 | 15 | 9 | 0.54 | 0.07 | -0.19 | 0.16 | 0.05 |
| ASXL1 | Endometrial Cancer | 78 | 7 | 0.26 | 0.0005 | 74 | 7 | 0.29 | 0.0014 | 0.03 | 0.16 | 0 |
| TET2 | Prostate Cancer | 73 | 37 | 1.8 | 0.0008 | 19 | 8 | 1.2 | 0.62 | -0.6 | 0.18 | 0.59 |
| ATM | Melanoma | 63 | 39 | 0.99 | 0.95 | 62 | 38 | 0.96 | 0.83 | -0.03 | 0.19 | 0 |
| ATM | Pancreatic Cancer | 97 | 69 | 0.74 | 0.02 | 80 | 55 | 0.7 | 0.0098 | -0.04 | 0.2 | 0.21 |
| ATM | Glioma | 65 | 50 | 0.95 | 0.72 | 63 | 48 | 0.93 | 0.6 | -0.02 | 0.22 | 0 |
| SF3B1 | Breast Cancer | 85 | 25 | 1.14 | 0.51 | 82 | 23 | 1.09 | 0.69 | -0.06 | 0.26 | 0 |
| ATM | Colorectal Cancer | 244 | 72 | 0.6 | 0 | 240 | 70 | 0.59 | 0 | -0.01 | 0.29 | 0 |
| SF3B1 | Bladder Cancer | 64 | 21 | 0.6 | 0.02 | 61 | 19 | 0.57 | 0.02 | -0.03 | 0.37 | 0.09 |
| ASXL1 | Glioma | 53 | 39 | 0.86 | 0.34 | 41 | 31 | 0.93 | 0.68 | 0.07 | 0.4 | 0 |
| ATM | Biliary Tract Cancer | 46 | 43 | 1.12 | 0.5 | 40 | 37 | 1.07 | 0.71 | -0.05 | 0.53 | 0 |
| ATM | Breast Cancer | 90 | 30 | 1.04 | 0.85 | 82 | 26 | 1.01 | 0.98 | -0.03 | 0.61 | 0 |
| DNMT3A | Breast Cancer | 220 | 66 | 0.98 | 0.86 | 39 | 10 | 0.86 | 0.64 | -0.12 | 0.66 | 0.03 |
| SF3B1 | Pancreatic Cancer | 58 | 39 | 0.81 | 0.19 | 45 | 31 | 0.78 | 0.18 | -0.03 | 0.76 | 0.04 |
| SF3B1 | Non-Small Cell Lung Cancer | 127 | 71 | 1 | 0.98 | 102 | 56 | 0.99 | 0.96 | -0.01 | 0.85 | 0 |
| ATM | Prostate Cancer | 73 | 26 | 1.24 | 0.29 | 64 | 22 | 1.25 | 0.31 | 0.01 | 0.87 | 0.01 |
| TET2 | Bladder Cancer | 64 | 30 | 0.9 | 0.56 | 37 | 15 | 0.87 | 0.6 | -0.02 | 0.88 | 0 |
| ASXL1 | Bladder Cancer | 62 | 21 | 0.63 | 0.04 | 49 | 16 | 0.64 | 0.08 | 0.01 | 0.91 | 0.67 |
| TET2 | Esophagogastric Cancer | 45 | 28 | 1 | 0.99 | 33 | 19 | 1.01 | 0.96 | 0.01 | 0.92 | 0 |
| TET2 | Pancreatic Cancer | 96 | 70 | 0.77 | 0.04 | 22 | 15 | 0.75 | 0.3 | -0.02 | 0.93 | 0.93 |
| DNMT3A | Pancreatic Cancer | 228 | 162 | 0.88 | 0.14 | 21 | 15 | 0.9 | 0.71 | 0.02 | 0.93 | 0.05 |
| DNMT3A | Non-Small Cell Lung Cancer | 482 | 276 | 0.97 | 0.61 | 97 | 48 | 0.97 | 0.84 | 0 | 0.98 | 0.01 |
| DNMT3A | Glioma | 133 | 87 | 0.66 | 0.0002 | 54 | 29 | 0.66 | 0.03 | 0 | 0.98 | 0.39 |
| TET2 | Non-Small Cell Lung Cancer | 216 | 124 | 0.99 | 0.94 | 90 | 48 | 0.99 | 0.97 | 0 | 0.99 | 0.01 |
| DNMT3A | Leukemia | 51 | 40 | 1.19 | 0.35 | 51 | 40 | 1.19 | 0.35 | 0 | 1 | 0 |
| SF3B1 | Melanoma | 41 | 25 | 0.99 | 0.95 | 41 | 25 | 0.99 | 0.95 | 0 | 1 | 0 |

#### Supplementary Table S8. Every affected gene-patient entry

All 12,009 gene-patient entries whose only evidence is a blood-normal specimen. The first 40 rows are shown here; the complete table is supplied as the tab-delimited file `CH_flags_to_drop.tsv` and as sheet S8 of the accompanying workbook. The study and sample_id columns identify the patient's representative tumor record, not the blood specimen that supplied the contaminating call.

| pk | gene | cancer_type | oncotree_code | study | sample_id | age |
|---|---|---|---|---|---|---|
| MSK:P-0000004 | TET2 | Breast Cancer | IDC | breast_msk_2018 | P-0000004-T01-IM3 | 39.7 |
| MSK:P-0000037 | DNMT3A | Hepatocellular Carcinoma | HCC | hcc_msk_2024 | P-0000037-T02-IM3 | 64.4 |
| MSK:P-0000039 | GNAS | Mesothelioma | PEMESO | msk_ch_2020 | P-0000039-N01 | 69 |
| MSK:P-0000059 | DNMT3A | Esophagogastric Cancer | EGC | msk_ch_2020 | P-0000059-N01 | 62.4 |
| MSK:P-0000062 | ASXL1 | Esophagogastric Cancer | GEJ | egc_msk_2017 | P-0000062-T01-IM3 | 79.4 |
| MSK:P-0000062 | TET2 | Esophagogastric Cancer | GEJ | egc_msk_2017 | P-0000062-T01-IM3 | 79.4 |
| MSK:P-0000066 | DNMT3A | Breast Cancer | BRCA | msk_ch_2020 | P-0000066-N01 | 61.9 |
| MSK:P-0000071 | TET2 | Bladder Cancer | BLCA | paired_bladder_2022 | P-0000071-T01-IM3 | 65.7 |
| MSK:P-0000081 | TET2 | Breast Cancer | IDC | breast_alpelisib_2020 | P-0000081-T01-IM3 | 50.3 |
| MSK:P-0000085 | SF3B1 | Bladder Cancer | BLAD | msk_ch_2020 | P-0000085-N01 | 74 |
| MSK:P-0000088 | ATM | Bladder Cancer | BLAD | msk_ch_2020 | P-0000088-N01 | 62.4 |
| MSK:P-0000095 | TSC2 | Germ Cell Tumor | NSGCT | gct_msk_2016 | P-0000095-T01-IM3 | 43.2 |
| MSK:P-0000096 | CHEK2 | Bladder Cancer | BLAD | msk_ch_2020 | P-0000096-N01 | 62.9 |
| MSK:P-0000098 | DNMT3A | Breast Cancer | IDC | msk_chord_2024 | P-0000098-T01-IM3 | 66.5 |
| MSK:P-0000106 | DNMT3A | Breast Cancer | BRCA | msk_ch_2020 | P-0000106-N01 | 54.6 |
| MSK:P-0000112 | CHEK2 | Esophagogastric Cancer | ESCA | egc_msk_2017 | P-0000112-T01-IM3 | 61.5 |
| MSK:P-0000117 | DNMT3A | Esophagogastric Cancer | STAD | egc_msk_2017 | P-0000117-T01-IM3 | 78.8 |
| MSK:P-0000120 | DNMT3A | Breast Cancer | IDC | msk_chord_2024 | P-0000120-T01-IM3 | 52.6 |
| MSK:P-0000120 | ERBB4 | Breast Cancer | IDC | msk_chord_2024 | P-0000120-T01-IM3 | 52.6 |
| MSK:P-0000131 | DNMT3A | Breast Cancer | BRCA | msk_ch_2020 | P-0000131-N01 | 65.7 |
| MSK:P-0000133 | ARID1B | Non-Small Cell Lung Cancer | LUAD | lung_msk_2017 | P-0000133-T01-IM3 | 81.6 |
| MSK:P-0000134 | DNMT3A | Gastrointestinal Stromal Tumor | GIST | gist_msk_2023 | P-0000134-T01-IM3 | 69.9 |
| MSK:P-0000136 | DNMT3A | Breast Cancer | IDC | breast_msk_2018 | P-0000136-T01-IM3 | 70.4 |
| MSK:P-0000138 | SETD2 | Breast Cancer | IDC | breast_alpelisib_2020 | P-0000138-T02-IM3 | 52.5 |
| MSK:P-0000141 | DNMT3A | Ovarian Cancer | CCBOV | msk_ch_2020 | P-0000141-N01 | 62.8 |
| MSK:P-0000153 | DNMT3A | Breast Cancer | ILC | breast_alpelisib_2020 | P-0000153-T02-IM3 | 53 |
| MSK:P-0000155 | DNMT3A | Breast Cancer | BRCA | msk_ch_2020 | P-0000155-N01 | 76.5 |
| MSK:P-0000165 | DNMT3A | Non-Small Cell Lung Cancer | LUAD | lung_msk_2017 | P-0000165-T01-IM3 | 66.7 |
| MSK:P-0000174 | ATM | Thyroid Cancer | THPD | thyroid_mskcc_2016 | P-0000174-T01-IM3 | 71 |
| MSK:P-0000174 | ATRX | Thyroid Cancer | THPD | thyroid_mskcc_2016 | P-0000174-T01-IM3 | 71 |
| MSK:P-0000174 | NOTCH1 | Thyroid Cancer | THPD | thyroid_mskcc_2016 | P-0000174-T01-IM3 | 71 |
| MSK:P-0000174 | TP53 | Thyroid Cancer | THPD | thyroid_mskcc_2016 | P-0000174-T01-IM3 | 71 |
| MSK:P-0000182 | CHEK2 | Hepatocellular Carcinoma | HCC | msk_ch_2020 | P-0000182-N01 | 72.2 |
| MSK:P-0000182 | DNMT3A | Hepatocellular Carcinoma | HCC | msk_ch_2020 | P-0000182-N01 | 72.2 |
| MSK:P-0000182 | EGFR | Hepatocellular Carcinoma | HCC | msk_ch_2020 | P-0000182-N01 | 72.2 |
| MSK:P-0000182 | TET2 | Hepatocellular Carcinoma | HCC | msk_ch_2020 | P-0000182-N01 | 72.2 |
| MSK:P-0000187 | DNMT3A | Breast Cancer | BRCA | msk_ch_2020 | P-0000187-N01 | 70.7 |
| MSK:P-0000199 | PIK3CA | Ovarian Cancer | CCBOV | msk_ch_2020 | P-0000199-N01 | 61.4 |
| MSK:P-0000206 | TET2 | Salivary Gland Cancer | SACA | msk_ch_2020 | P-0000206-N01 | 82.7 |
| MSK:P-0000207 | POLE | Breast Cancer | ILC | breast_msk_2018 | P-0000207-T01-IM3 | 54.9 |

S8 lists every gene-patient entry set by a call arising in a blood-normal specimen. Removing these entries from a patient-keyed alteration matrix reproduces the corrected matrix analysed in this report. cancer_type = "Unknown or unassignable primary" (441 rows, 353 patients) denotes a record whose contributing study labelled the primary site "Unknown" or "Cancer of Unknown Primary", or gave a hepatobiliary umbrella label that could not be resolved to a specific OncoTree code. These patients are excluded from the solid-tumor analysis set but their flagged entries are listed here, because a matrix built without that exclusion still contains them. age is age in years at sequencing, rounded to one decimal place. sample_id and study are reproduced verbatim from the public cBioPortal deposits; no field in this file contains information that is not already retrievable from the portal.
